# Partial Prediction of the Virus COVID-19 Spread in Russia Based on SIR and SEIR Models

**DOI:** 10.1101/2020.07.05.20146969

**Authors:** Dmitry Tomchin, Alexander Fradkov

**Affiliations:** Institute for Problems of Mechanical Engineering, Saint Petersburg

## Abstract

The possibility to predict the spread of COVID-19 in Russia is studied. Particular goal is to predict the time instant when the number of infected achieves its maximum (peak). Such a partial prediction allows one to use simple epidemoics models: SIR and SEIR. Simplicity and small number of parameters are significant advantages of SIR and SEIR models under conditions of a lack of numerical initial data and structural incompleteness of models. The prediction is carried out according to public WHO datasets from March 10 to April 20, 2020. Comparison of forecast results by SIR and SEIR models are given. In both cases, the peak number of infected persons while maintaining the current level of quarantine measures is forecasted at the end of May 2020 or later. It coincides with the real data obtained in May-June, 2020. The results confirm usefulness of simple nonlinear dynamical models for partial prediction of complex epidemic processes.

## 1 Introduction

The pandemic of the new coronavirus COVID-19 lead to appearance of numerous new research problems. One of them is prediction of the epidemics spread with a reasonable accuracy based on limited amount of data [1]. Such a prediction is needed, e.g. for prediction of the lockdown and quarantine measures efficiency. In a complex, difficult to predict situation, an efficient approach to prediction is based on using mathematical models of epidemics. A number of approaches are used for epidemics prediction. Some of them use sophisticated data models of higher dynamical order [2, 3]. In order to increase the accuracy of such models data from a longer time interval are needed. The other results are based on on first principles dynamical models and their modifications. The basic epidemics models have been known for over a century and still attract the attention of specialists. In a number of papers the classical models SIR, SEIR and their modifications [5]

In this paper, an attempt is made to use the simplest epidemic models: SIR and SEIR to predict the spread of COVID-19 in Russia. Despite their simplicity, they are often used in epidemiology. They were used for the analysis of the current epidemic of the coronavirus COVID-19 [6, 7, 8]. For example, on April 22, 2020, out of 598 publications in ArXiv.org, the title or abstract of which contains the words COVID-19 or SARS-CoV-2, 32 have the term SIR, and 12 have the term SEIR. ^1^ In some works the advantages of a simpler SIR model over a more complex model SEIR are mentioned [11]. Simplicity and a small number of parameters are the advantages of both SIR and SEIR models, which are very significant under conditions of a lack of numerical initial data and structural incompleteness of the models. SIR and SEIR models were used to predict the distribution of COVID-19 in China, France, Italy, Germany, Portugal and several other countries [6, 7, 8, 9, 10] Some results on forecasting the distribution of COVID-19 in Russia as a whole are available in [7].

In this work, we attemplt to forecast the spread of COVID-19 in Russia over a more representative dataset and compare the results of the forecast for SIR and SEIR models based on official statistics on incidence. To set the parameters of the models, which are difficult to determine from the official data, a scenario approach is used: the dynamics of the epidemic is analyzed at several possible parameter values. Some of the SEIR model parameters are selected using the [7] results.

## 2 Forecasting by SIR Model

In the classical SIR model (Kermack-McKendrick model) [12] three groups of individuals are considered: susceptible (S), infected (I) and recovered/removed (R). Transmission of infection occurs from infected individuals to susceptible ones. It is believed that the recovered individuals acquire immunity and cannot be infected for the second time. The model is described by differential equations

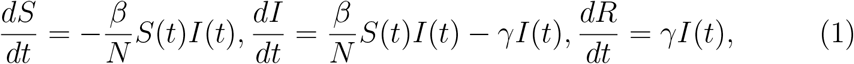

where *S*(*t*), *I*(*t*), *R*(*t*) is the numbers of susceptible, infected and recovered, correspondingly. Positive numbers *β, γ* are interpreted as parameters defining the rate of infection and the rate of recovering, correspondingly. Details see in [13].

To apply the model to a specific situation, it is necessary to calibrate it i.e. to determine the values of the parameters and the initial conditions from the experimental data. We will use the official data of the World Health Organization (WHO), presented in a convenient form on the website Worldometers [14]. Since WHO data is updated once a day, it is convenient to switch from a differential model (1) to a discrete one:

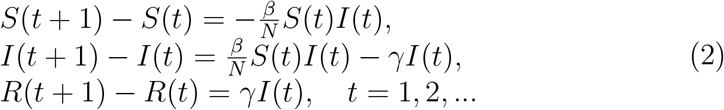

In our study the [14] data for calibration from March 10, 2020 to April 20, 2020 were used for calibration. The initial conditions are equal to the values of the variables on March 10, 2020.

To estimate the parameters, we use the least squares method. We apply it in two steps: first evaluate *γ* using the formula

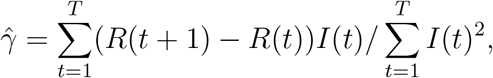

and then estimate *β* from the expression

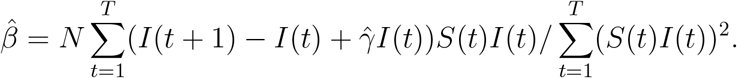

Let *N* be the initial number of susceptible coinciding with the population of the Russian Federation: *N* = 146.745 million. The calculation yields the following parameter values: 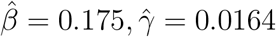.

Model calibration results are shown in Fig.1. At first glance, the error seems to be large, but it must be kept in mind that the model should first of all take into account the total number of infected people, rather than the rate of its growth. However, one can try to refine the estimates. From the graphs it can be seen that the accuracy of the approximation decreases after March 27, since the real rate of increase in incidence decreases. Obviously, this is due to the introduction of partial self-isolation in the Russian Federation during the third decade of March. For a more accurate prediction, taking into account self-isolation, the time interval of observations was divided into two parts: 1) before March 28 and 2) from March 28 to April 20 and the model parameters on each set were evaluated separately. If we evaluate the parameters of the model separately from the data from March 11 to 20, we get 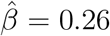, while according to the data from March 21 to April 15 we get 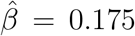, i.e. the same value as in the first case. The estimates of the parameter *γ* also coincide in both cases: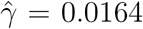. The results of model calibration using a truncated data set are presented in Fig.2.

**Figure 1:**
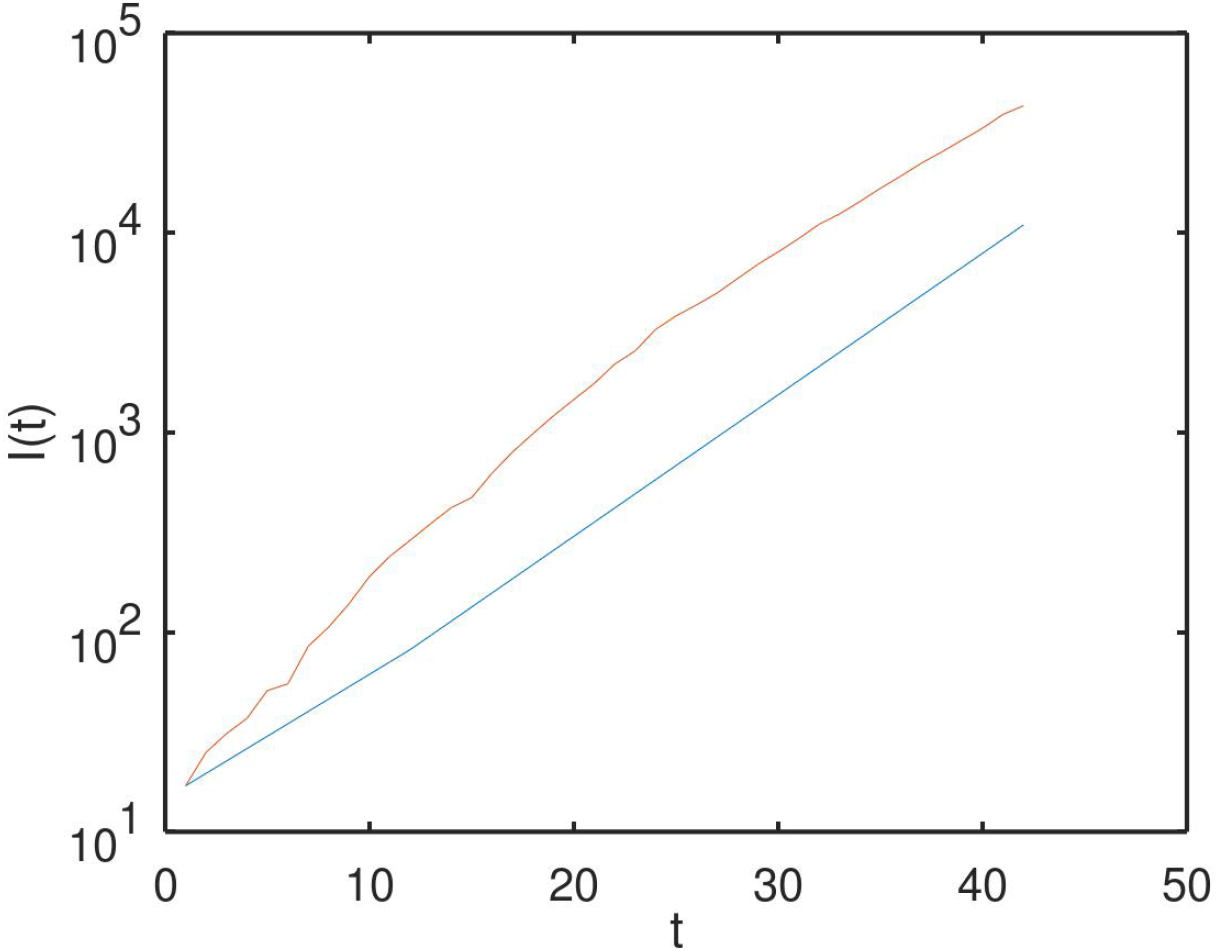
Calibration of the SIR model according to the data from 2020/03/10 to 2020/04/20. The red line corresponds to the real data, the blue line is the graph of the variable *I*(*t*) according to the model.

**Figure 2:**
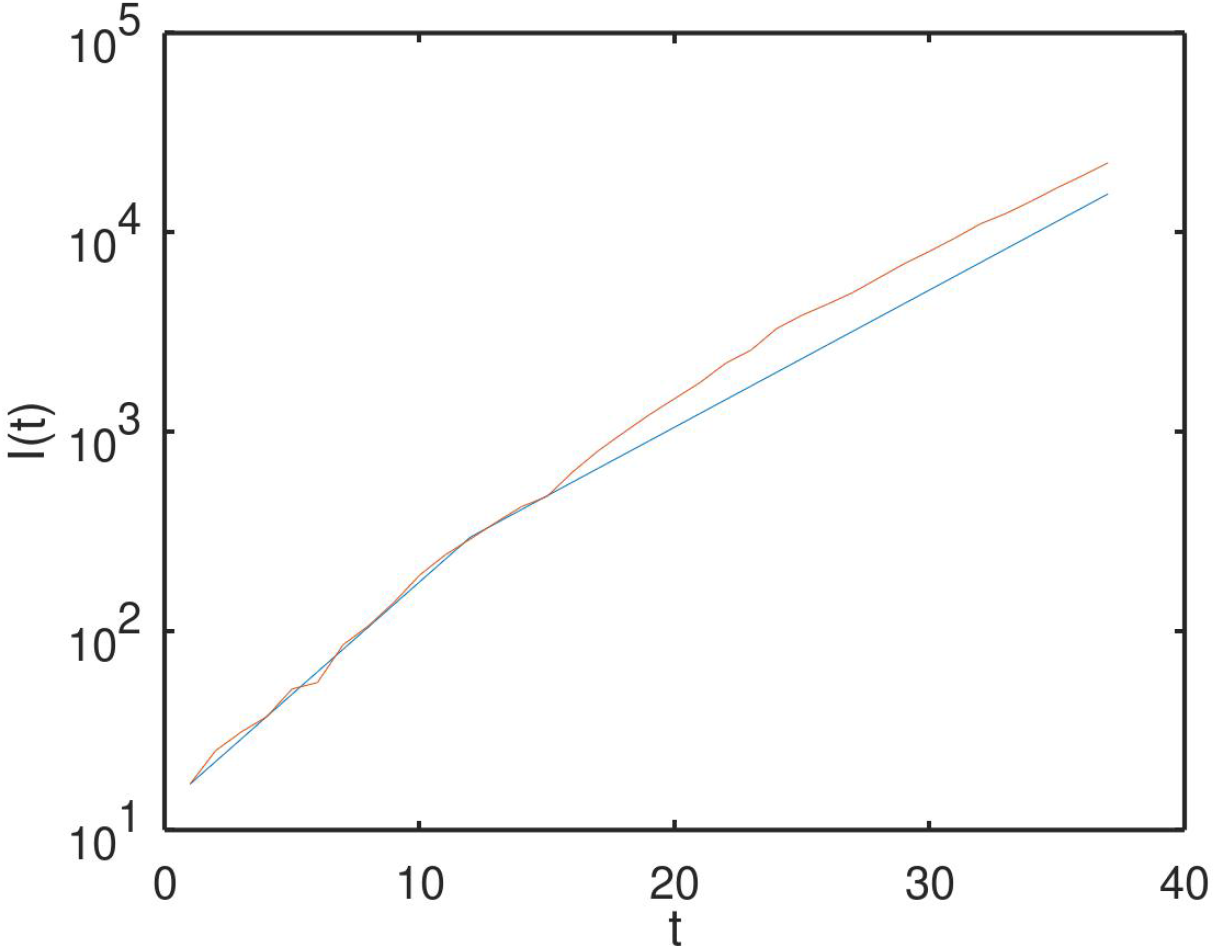
Calibration of the SIR model separately according to data from 10.03.2020 to 03.27.2020 and from 28.03.2020 to 04.16.2020. The red line corresponds to the real data, the blue line is the graph of the variable *I*(*t*) according to the updated model.

The results of the forecast of the development of the epidemic 120 days in advance are presented in Fig. 3. In Fig. 3, a forecast for the period after April 20 is based on data from March 10 till April 20. The graph shows that the peak incidence (reaching a plateau) is predicted to be around the 70th day, i.e. June 30, 2020. For comparison, in Fig. 3, b, for the forecast for the period after March 28, the values ??of the model parameters calculated from the data of March 1028, i.e. before the introduction of self-isolation are used. The graph shows that if the self-isolation mode were not introduced, the peak would have come earlier: on the 60th day, i.e. 26 of May and its height would be significantly higher. The results can be used to assess the intensity of quarantine measures required for a given slowdown in the development of outbreaks of epidemics and pandemics.

**Figure 3:**
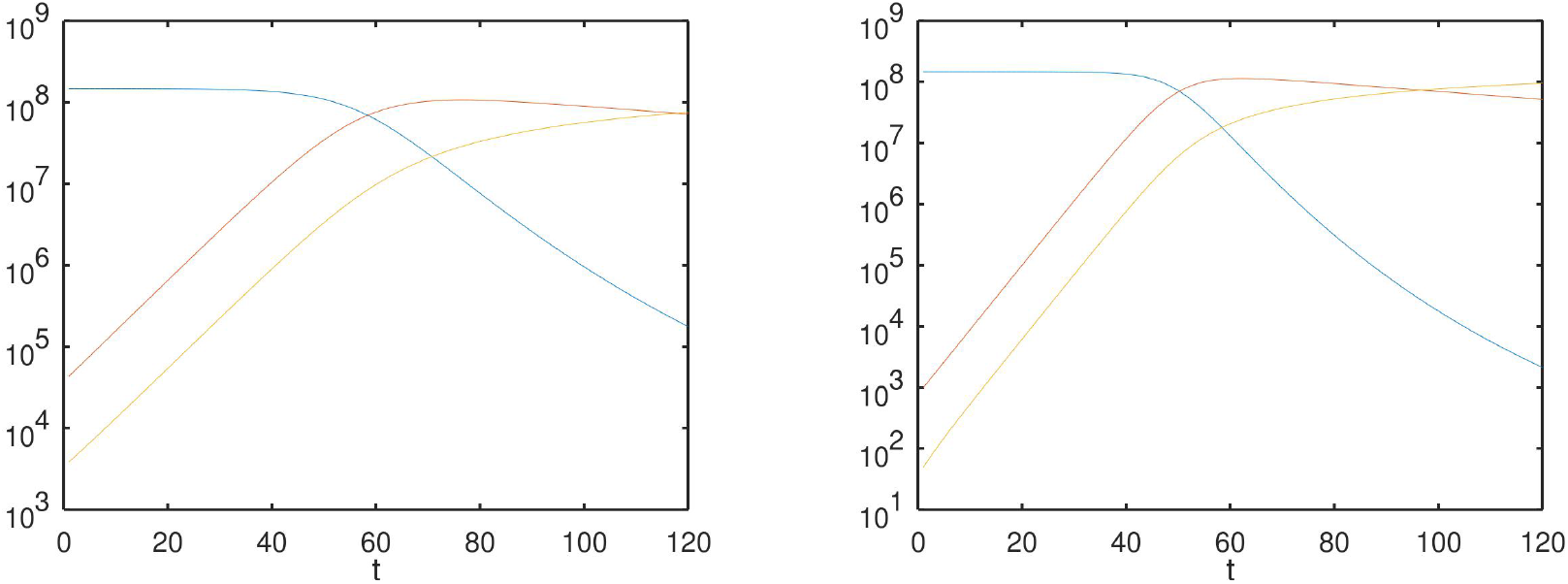
Forecast from SIR model (2) for 120 days a) (left): from Apr. 20 till Aug. 18, 2020 based on the data from March, 10 till April, 20; b) (right): from March 27 till July 25, 2020 based on the data from March 11 till March 20. Blue line corresponds to the number of susceptible *S*(*t*); red line is the number of infected *I*(*t*), yellow line is the number of removed *R*(*t*) (sum of the number of recovered and the number of the deseased).

Thus, even the simplest SIR model shows the efficiency of introducing a self-isolation regime from the point of view of stretching the incidence growth phase over time and allows us to give useful estimates of the incidence growth time. As for the number of infected people, its prediction at first glance looks disappointing: in order to begin to decrease, this number should approach the total population of the country. However, it should be kept in mind that in the population there are a large number of asymptomatic infected people who easily tolerate the disease often do not even knowing about it. The ability to quantify asymptomatic patients will be shown in the next section using the SEIR model.

## 3 Forecasting by SEIR Model

The SEIR model (Susceptible Exposed Infected Recovered) takes into account the incubation period of the disease [13]. This model was used to predict the spread of COVID-19 in China [6] and in European countries [7]. We will use the modified SEIR model with discrete time [7], described by the equations

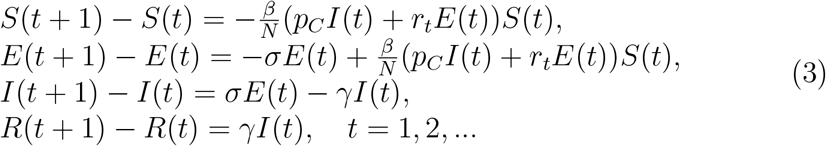

Here *S*(*t*), *I*(*t*), *R*(*t*) have the same meaning as in the SIR model (2), and *E*(*t*) is the number of the infected individuals in the stage of incubation (latent) period. Total population *N* = *S*(*t*) + *E*(*t*) + *I*(*t*) + *R*(*t*), is assumed to be constant as before.

The SEIR model (3) has two parameters the values ??of which can be estimated based on real statistical data: *γ, β*. The parameter *γ >* 0 represents the intensity of mortality and recovery, the parameter *β >* 0 corresponds to the rate of infection of the virus by susceptible people during contact with infected or latent ones. The parameter *σ >* 0 determines the incubation rate at which symptoms appear in individuals in the latent period. The value of *σ* can be chosen inversely proportional to the average incubation period of COVID-19: *σ* = 1*/*7.

The number *p*_*C*_ *>* 0 corresponds to the number of contacts per person per day for infected *I*(*t*) (it is assumed that if infected people with symptoms are quarantined (lockdown) then the number of contacts decreases); *r*_*t*_ *> p*_*C*_ is the number of contacts per person per day for the latent period *E*(*t*). Parameters *r*_*t*_, *p*_*C*_ can be changed by applying measures that regulate social distance (quarantine, lockdown). They also depend on population density and social traditions.

An important peculiarity of the COVID-19 pandemic, as well as a number of previous epidemics, is the significant difference between the actual number of infected individuals and the number of the documented ones. This is due to the presence of a significant number of asymptomatic infections, due to the inability to carry out full testing, as well as due to the inaccuracy and insufficient sensitivity of the tests. Statistical data for Europe and the United States suggest that the proportion of undocumented cases can vary from 40% to 90% [15]. Denote by *α* the ratio of the total number of infected to the number of documented infected. Given the significant uncertainty of *α* and the difficulty of determining *α*, we will carry out calculations for *α* = 5 ^2^ and for *α* = 10.

First consider the case *α* = 5. As a result of evaluating the parameters of the SEIR model by LS method according to data from March 10 to April 20, we get: *β* = 0.027, *γ* = 0.017. Following [7] and based on the proximity of social traditions in France and in Russia, we will adopt the following values ??of the remaining parameters as basic ones: *σ* = 1*/*7; *p*_*C*_ = 2, *r* = 10. The above values ??correspond to the current quarantine mode (model-1). The initial conditions *S*(*t*), *I*(*t*), *E*(*t*), *R*(*t*) for *t* = 0 are also set similarly to [7], using the relations *E*(*t*) = (*I*(*t* + 1) *−* (1 *− γ*)*I*(*t*)*/σ, S*(0) = *N, I*(*t*) = *α*(*Ī*(*t*) *− D*(*t*) *− H*(*t*)), *R*(*t*) = *D*(*t*) + *αH*(*t*). The data on the number of infected *Ī*(*t*), the number of recovered *H*(*t*) and the number of dead *D*(*t*) are taken from official sources [14]. Calibration results are presented in Fig. 4. They show reasonable accuracy of approximation of real data by the model.

**Figure 4:**
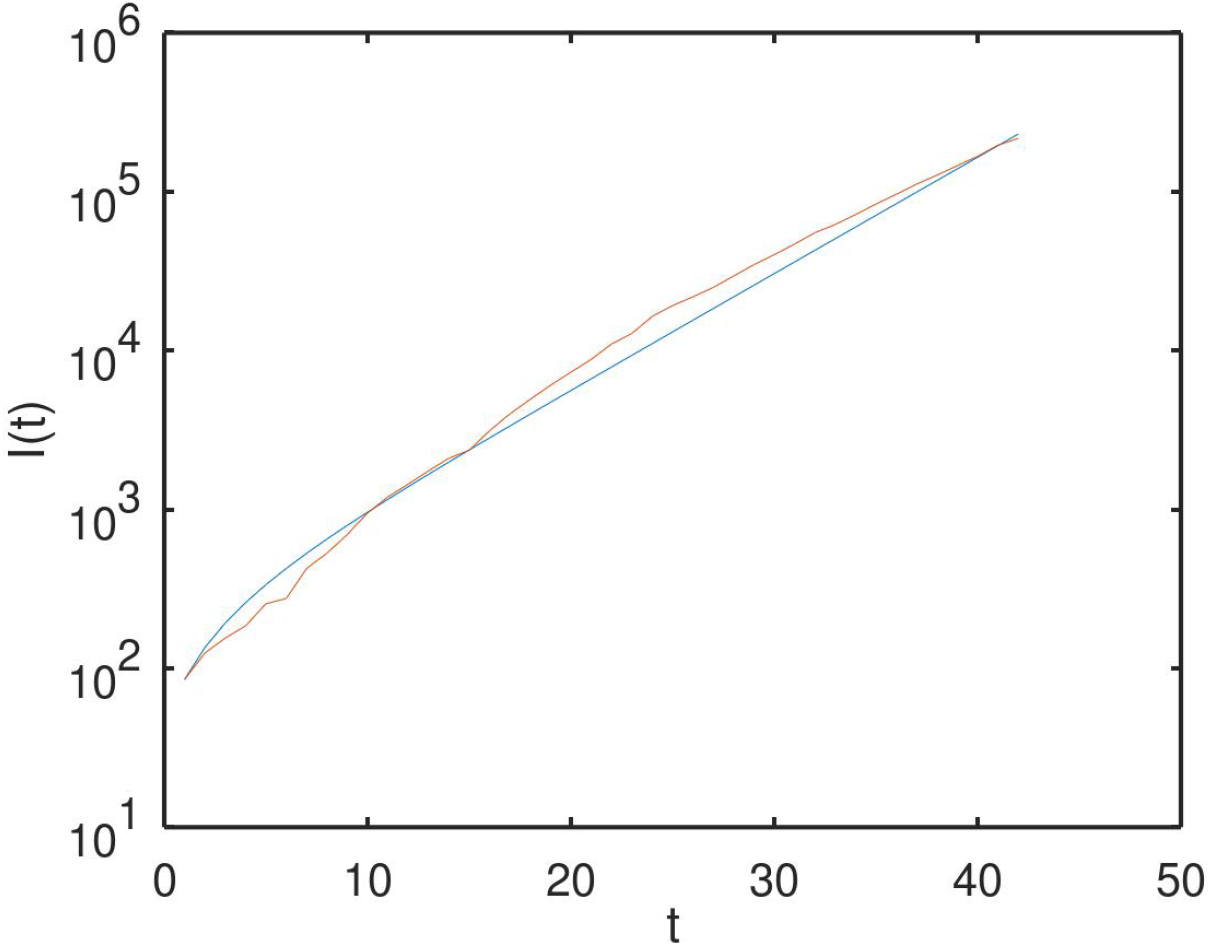
Calibration of the SEIR model at *α* = 5 according to data from 10.03.2020 till 04.20.2020. The blue line corresponds to the number of infected evaluated from the model, while the red line corresponds to the real data *Ī*(*t*).

The forecast results for 120 days according to the model are presented in Fig. 5. In Fig. 5a) (left) the results of the forecast for the period from April 20 to August 18, 2020 according to data from 10.03.2020 to 20.04.2020 are shown. In the graphs, the blue curves correspond to the function *S*(*t*), the yellow curves correspond to the function *E*(*t*), the red curves correspond to the function *I*(*t*), and the violet curves to the function *R*(*t*). It can be seen that the peak of the epidemic is predicted on the 40th day that is May 30, 2020.

**Figure 5:**
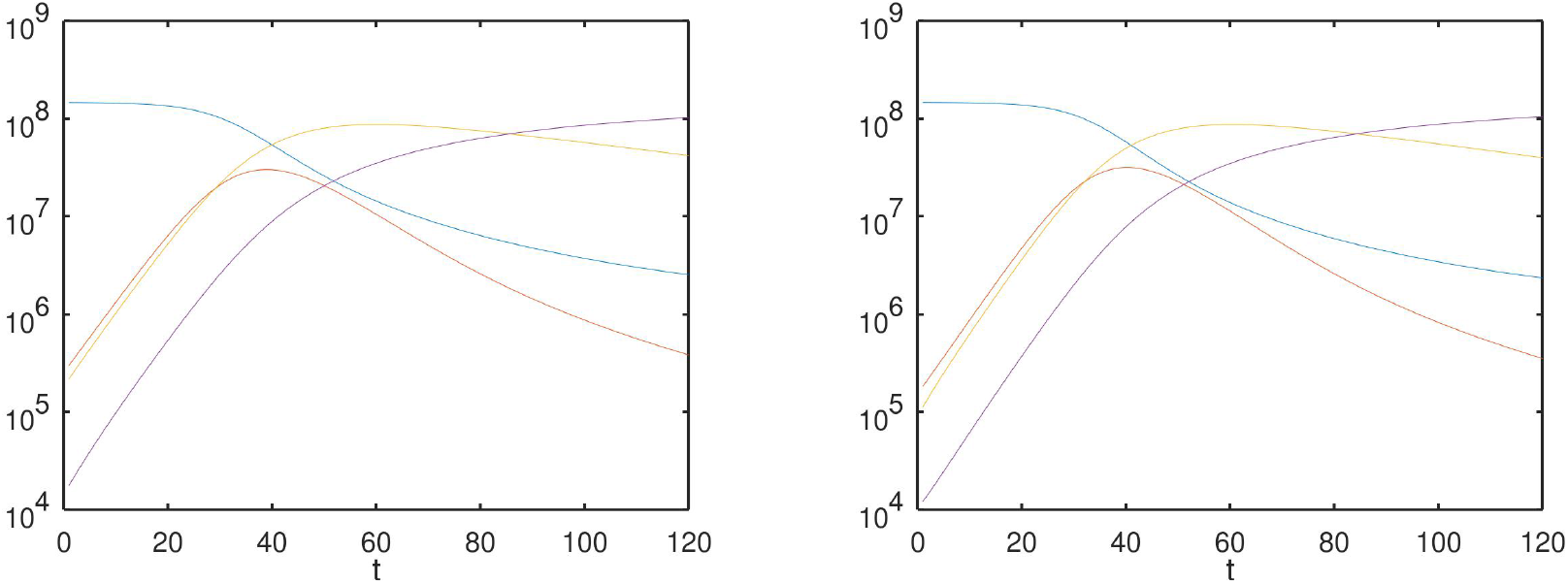
Forecast based on SEIR model-1 for 120 days at *α* = 5: a) (left) forecast from April 20 till August 18, 2020 according to data from 03.10.2020 till 04.20.2020. b) (right) - forecast from April 15 till August 13, 2020 according to the data from 10.03.2020 till 15.04.2020. The blue line is the number of susceptible *S*(*t*); the yellow line is the number of infected in the incubation period *E*(*t*), the red line is the number of infected *I*(*t*), the violet line is the number of removed *R*(*t*).

For comparison, in Fig. 5, b) the results of the forecast for the model built according to the data from March 10 to April 15 are shown. In such a model less data are used and it would seem that it should have less predictive power. However, the figure shows that even such an “outdated” model predicts the peak of the number of infected individuals with approximately same accuracy: on the 45th day or also on May 30th.

In Fig. 6 a similar forecast for the enhanced quarantine mode (model-2: *p*_*C*_ = 1.5, *r* = 7.5) is presented. In Fig. 7 the forecast for an even more strict quarantine mode (model-3: *p*_*C*_ = 1, *r* = 5) is presented. Thus, as can be seen from Figs. 5-7, in the standard quarantine mode, the peak incidence occurs on the 40th day - May 30, 2020, in strict mode - on the 60th day - June 10, and in the very strict mode on the 100th day - July 20, 2020. It is also seen that a more strict regime leads to a decrease in the peak incidence rate: in the standard regime, the number of cases can reach 50 million, in the strict quarantine regime the magnitude of the peak drops to 20 million, and in a very strict regime it becomes less than 10 million. Note that the number of documented cases is still *α* times less.

**Figure 6:**
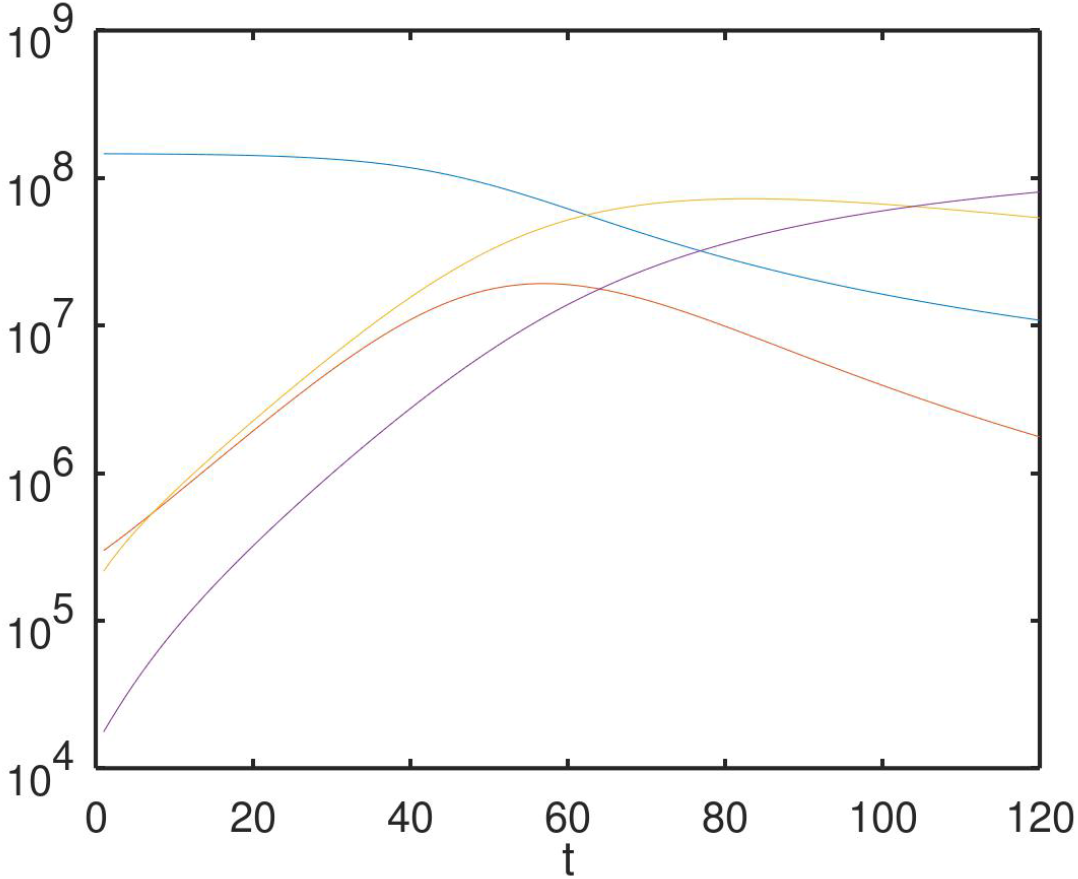
Forecast based on SEIR model-2 (strict quarantine) for *α* = 5 according to data from 10.03.2020 till 20.04.2020.

**Figure 7:**
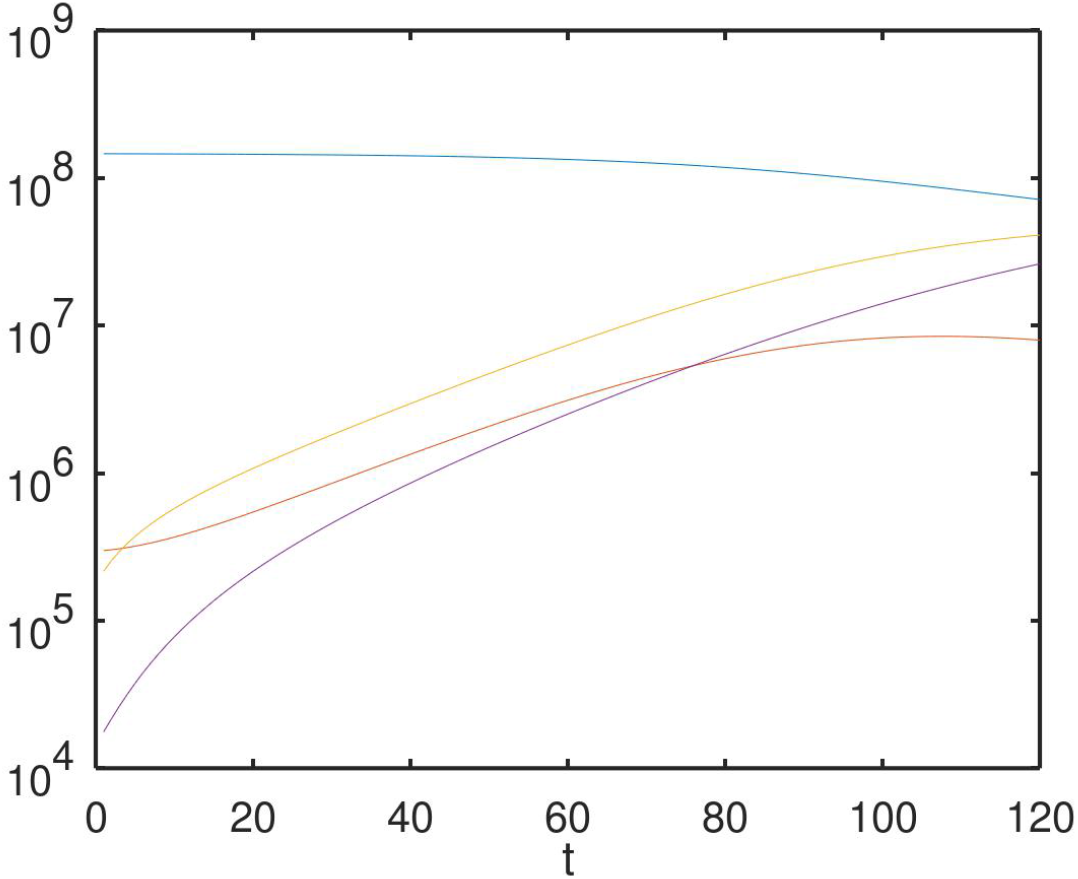
Forecast based on SEIR model-3 (very strict quarantine) at *α* = 5 according to data from 10.03.2020 till 04.20.2020.

We proceed to the case *α* = 10. The model is calibrated similarly to the case *α* = 5. Calibration results are presented in Fig.8. They exhibit good accuracy of approximation of real data by the model everywhere, except for the initial section. The forecast results for 120 days for the period from April 20 till August 18, 2020 according to the model are presented in Fig. 9. The graphs show that the predicted dates for reaching the maximum number of infected people will fall on May 23-24.

**Figure 8:**
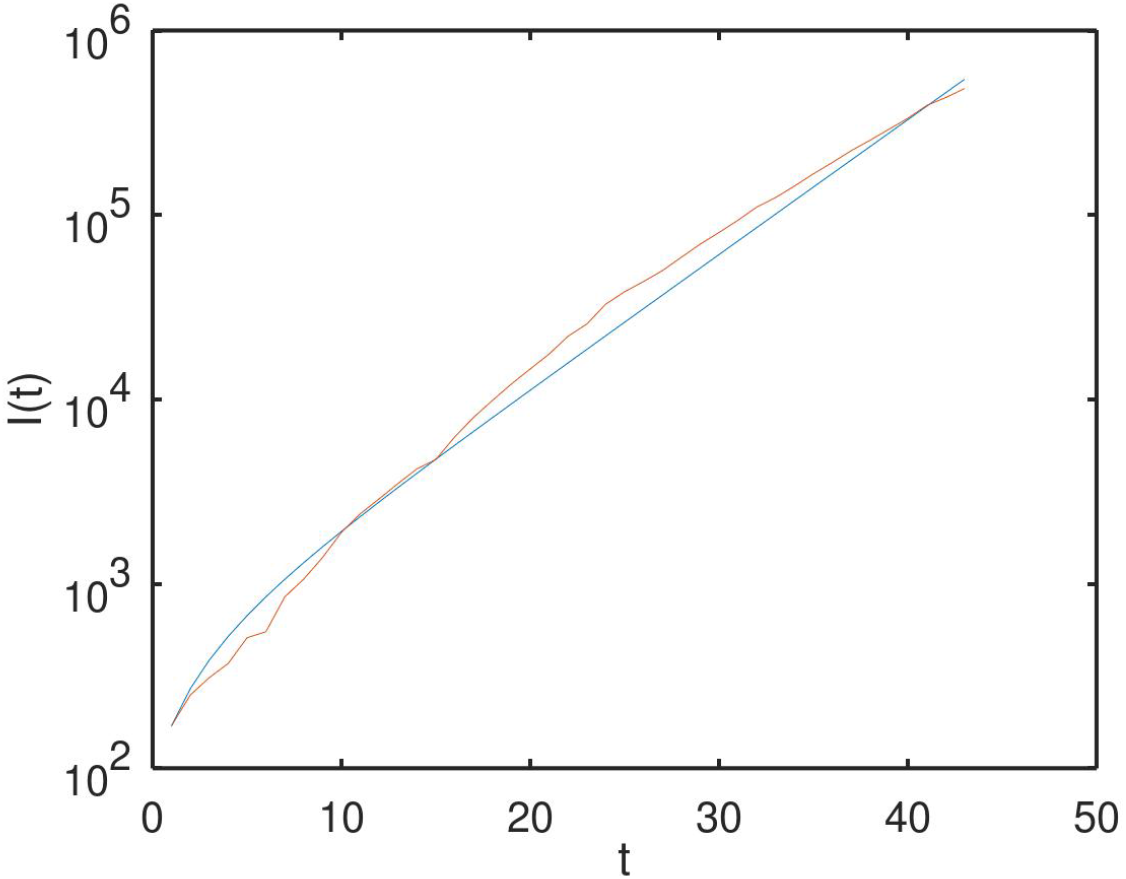
Calibration of the SEIR model at *α* = 10 according to data from 03.10.2020 till 04.20.2020. The blue line corresponds to the number of infected by the model, and the red line corresponds to the real data *Ī*(*t*).

**Figure 9:**
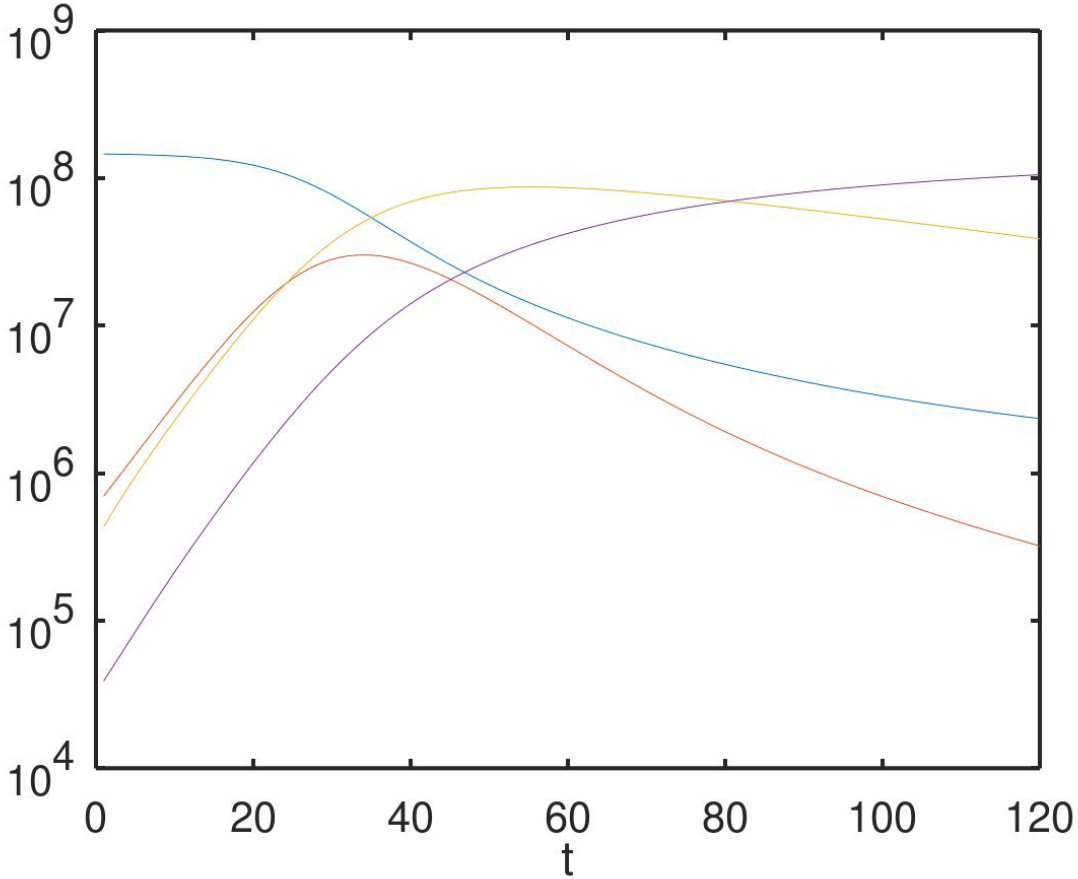
Forecast based on SEIR model-1 for 120 days ahead at *α* = 10: a) (left) forecast from April 20 till August 18, 2020 according to data from March 10, 2020 - April 20, 2020. b) (right) - forecast from April 15 till August 13, 2020 according to the data of 10.03.2020 - 15.04.2020. The blue line is the number of susceptible *S*(*t*); yellow line is the number of infected in the incubation period *E*(*t*); the red line is the number of infected *I*(*t*); the violet line is the number of removed *R*(*t*).

In Fig. 10 a similar forecast for strict quarantine mode (model-2, *p*_*C*_ = 1.5, *r* = 7.5) is presented. Fig. 11 demonstrates forecast for a “very strict “quarantine mode (model-3: *p*_*C*_ = 1, *r* = 5). It can be seen that the change in the behavior of the processes with a change in the value of *α* from *α* = 5 to *α* = 10 is not significant.

**Figure 10:**
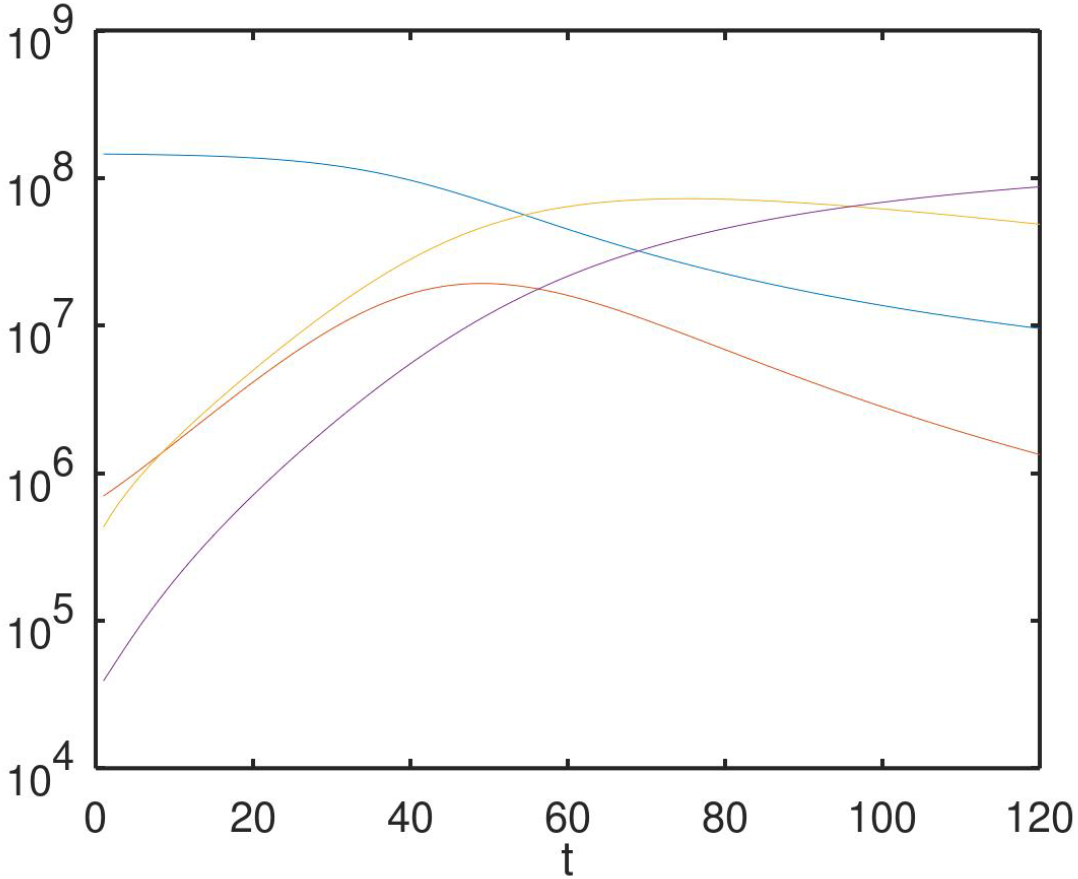
Forecast for SEIR model-2 (strict quarantine) for *α* = 10 according to the data of 10.03.2020 - 04.20.2020.

**Figure 11:**
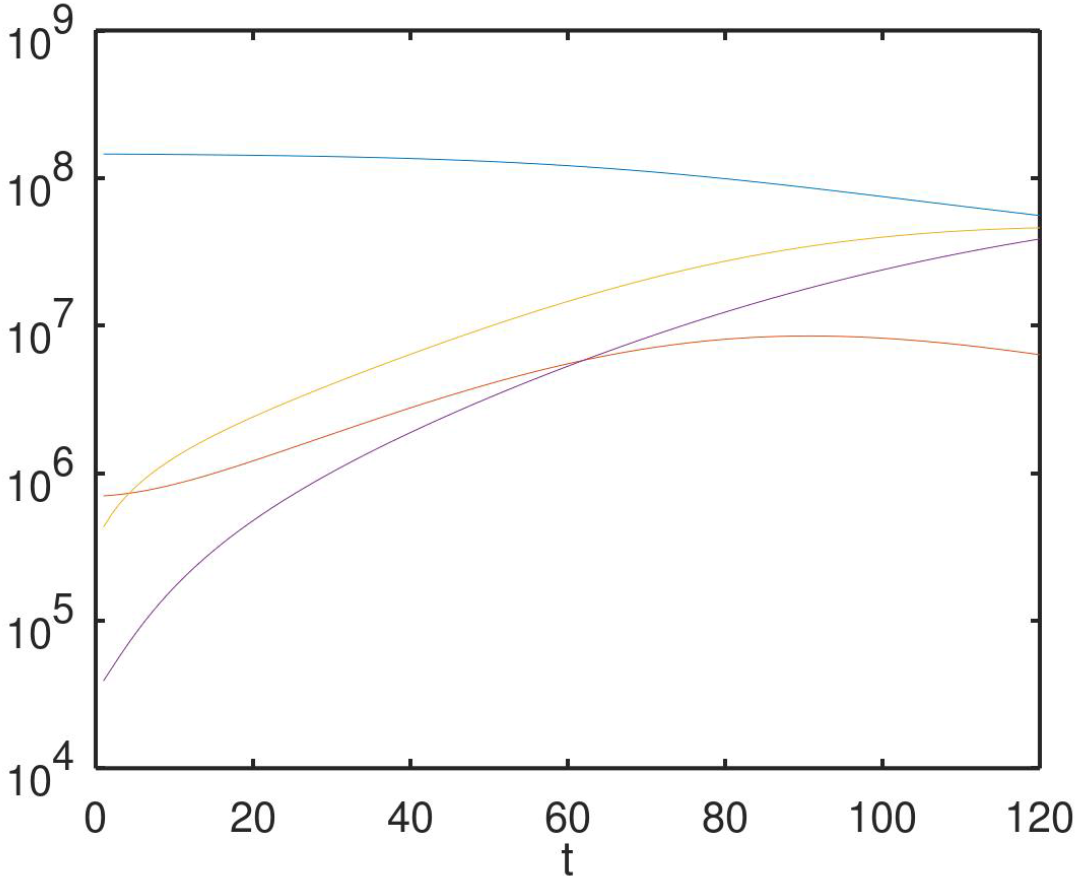
Forecast for a “very strict “quarantine mode (model-3: *p*_*C*_ = 1, *r* = 5) for *α* = 10 according to the data of 10.03.2020 - 04.20.2020.

## 4 Conclusion

In the paper the mathematical modelling of the spread of the COVID-19 Coronavirus Disease Based on Statistics from incidence in Russia from March 10 to April 20, 2020 is discussed. Two classical simple models of epidemiology are considered: SIR and SEIR. The parameters of the models are estimated based on published statistical data. Options for predicting the development of morbidity in the Russian Federation for 120 days are built when changing the parameter of social distance corresponding to different modes of isolation of the population (quarantine) and the parameter *α*, which determines the proportion of undocumented cases of infection.

Our calculations show that the SIR and SEIR models can produce reasonable results under conditions of limited amount of data and lack of reliability of statistics. They allow one to obtain not only qualitative, but also quantitative forecasts of the spread of the virus. In particular, it follows from the calculations that the peak of the number of infected in Russia should be revived no earlier than the end of May. The real WHO data, see Fig.12 [14] confirm the above conclusions. The forecast results can be used to evaluate the efficiency of lockdown regimes, to select the optimal strategy, for example, by periodically strengthening and weakening quarantine measures [16], etc.

**Figure 12:**
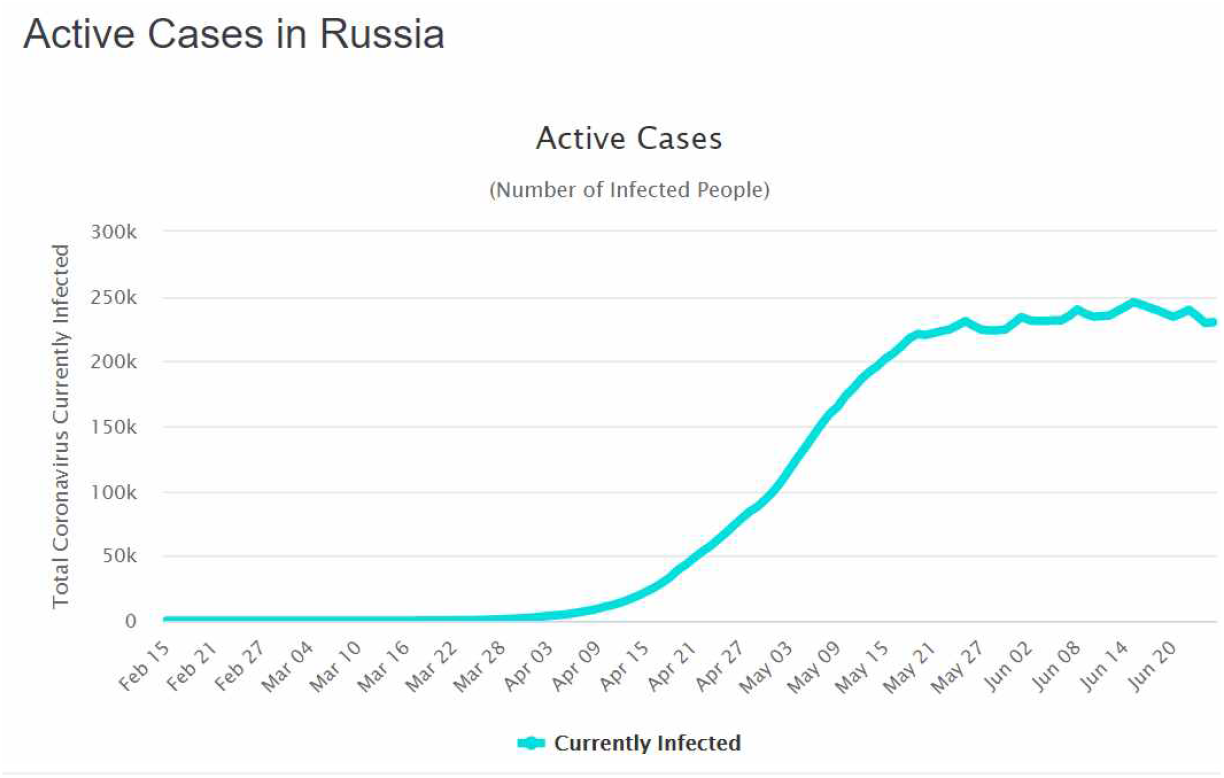
Number of infected in Russia from Feb.15, 2020 till June 25, 2020 according to [14]

It is worth to note that in April-May, 2020, the media repeatedly expressed, with reference to officials, various statements about time of the onset of the “peak” or “plateau” of the spread of the COVID-19 virus in Russia. In mid-April they spoke of a peak in late April or early May 2020, and in late April and early May they spoke of a peak in mid-May. See, e.g. the following quotations.

“MOSCOW, April 7. / TASS /. The situation with the spread of coronavirus in Russia will reach a plateau after about 10-14 days, the head of the Federal Biomedical Agency said Veronika Skvortsova on Tuesday in an interview with Russia-24” [17].

“MOSCOW, 29 Apr RIA Novosti. Sberbank using artificial intelligence have built a model that predicted the timing of the peak of the epidemic of coronavirus in Russia: across the country he will be in the first decade of may and in Moscow there will come a few days earlier, said the first Deputy Chairman of Board of Bank Alexander Vedyahin in an interview with RIA Novosti”[18].

However, the forecast results from the SEIR model based on the data from March 10 to April 15, show that such an “outdated” model predicts that the peak of the number of infected may happen on the 45th day, i.e. at the end of May. In the end of June, 2020 one can see that our forecasts of the peak dates are closer to reality than the official ones. References to the use of artificial intelligence in models without detailed explanations [18] are more likely to be suspicious than credible.

It is worth noticing that the calculations were carried out under a number of additional assumptions: it was believed that the entire population of the country was susceptible to the virus, the virus does not have seasonality, etc. Under such conditions, the final scenario is that a significant part of the population will eventually become ill, only the rate of spread of the disease will change. In reality, the spread of infection can be limited by some factors that are currently unknown and were not taken into account here.

Further studies should be carried out for models that take into account spatial heterogeneities (network or compartment models) and incomplete susceptibility of the population to infection. In addition it is interesting to compare our forecasting results with those obtained from modified SIR/SEIR models, e.g. with the results based on the fSIR (feedback SIR) model [5] or other simple models, e.g. ARIMA [3].

## Data Availability

Only freely available WHO data on COVID-19 spread are used in the paper.

https://www.worldometers.info/coronavirus/country/russia/

## Footnotes

1 Note, that the publication of six hundred articles on the problem in just two months indicates the serious interest of researchers in this topic. And in two more months, on June, 21 there were already 1,382 papers related to COVID-19 in Arxiv.

2 Similar values ??were also taken in studies of the development of the COVID-19 pandemic in France [7]

## References

[1] Boccaletti S., Ditto W., Mindlin G., Atangana A. Modeling and forecasting of epidemic spreading: The case of Covid-19 and beyond Chaos, Solitons and Fractals 135 (2020) 109794.

[2] Srivastava A., Prasanna V.K. Learning to Forecast and Forecasting to Learn from the COVID-19 Pandemic arXiv:2004.11372v3 [q-bio.PE] 4 May 2020.

[3] Perone G. ARIMA forecasting of COVID-19 incidence in Italy, Russia, and the USA arXiv:2006.01754 [statAP] 5 June, 2020.

[4] Postnikov E.B. Estimation of COVID-19 dynamics on a back-of-envelope: Does the simplest SIR model provide quantitative parameters and predictions? Chaos, Solitons and Fractals 135 (2020) 109841.

[5] Franco E. A feedback SIR (fSIR) model highlights advantages and limitations of infection-based social distancing. arXiv:2004.13216v2 [q-bio.PE] 4 May 2020.

[6] Z. Yang, Z. Zeng, K. Wang, S.-S. Wong, W. Liang, M. Zanin, P. Liu, X. Cao, Z. Gao, Z. Mai, J. Liang, X. Liu, S. Li, Y. Li, F. Ye, W. Guan, Y. Yang, F. Li, S. Luo, Y. Xie, B. Liu, Z. Wang, S. Zhang, Y. Wang, N. Zhong, and J. He, Modified SEIR and AI prediction of the epidemics trend of COVID-19 in China under public health interventions, Journal of Thoracic Disease, vol. 12, no. 3, 2020. [Online]. Available: http://jtd.amegroups.com/article/view/36385 xI, I, 1, I, II, III, 3, III, 4, IV-B, IV-C

[7] Efimov D., Ushirobira R. On an interval prediction of COVID-19 development based on a SEIR epidemic model. [Research Report] INRIA. 2020, ffhal-02517866v4f

[8] Zhong L., Mu L., Li J., Wang J., Yin Z., Liu D. Early Prediction of the 2019 Novel Coronavirus Outbreak in the Mainland China Based on Simple Mathematical Model. IEEE Access, V.8, 51761–51769, March 24, 2020, DOI: 10.1109/ACCESS.2020.2979599

[9] Teles P. A time-dependent SEIR model to analyse the evolution of the SARS-CoV-2 epidemic outbreak in Portugal arXiv:2004.04735, April 21, 2020.

[10] The Institute for Health Metrics and Evaluation (IHME). http://www.healthdata.org/covid

[11] Roda, W.C., Varughese, M.B., Han, D., Li, M.Y. Why is it difficult to accurately predict the COVID-19 epidemic? (2020) Infectious Disease Modelling, 5, pp. 271–281.

[12] Kermack W. O., McKendrick, A. G. A Contribution to the Mathematical Theory of Epidemics // Proc. Roy. Soc. Lond., Ser. A. 1927. No. 115. pp. 700721.

[13] Wang, Z.; Bauch, C.T.; Bhattacharyya, S. et al. Statistical physics of vaccination. Physics Reports, Vol. 664, pp.1–113, 2016.

[14] Worldometers. https://www.worldometers.info/coronavirus/country/russia/

[15] Li R., Pei S., Chen B., Song Y., Zhang T., Yang W., Shaman J., Substantial undocumented infection facilitates the rapid dissemination of novel coronavirus (SARS-CoV2). Science, 16 Mar 2020, 10.1126/science.abb3221 (2020).

[16] Eubank S., Eckstrand I., Lewis D., Venkatramanan S., Marathe M., Barrett C.L. Commentary on Ferguson, et al., Impact of Nonpharmaceutical Interventions (NPIs) to Reduce COVID-19 Mortality and Healthcare Demand Bull Math Biol. 2020; 82(4): 52. Published online 2020 Apr 8. doi: 10.1007/s11538-020-00726-x

[17] RBC: Apr. 7, Skvortsova announced the remaining 10-14 days before the plateau of the coronavirus. https://www.corona24.news/c/2020/04/07/skvortsova-announced-the-remaining-10-14-days-before-the-plateau-of-the-coronavirus-society-rbc-2.html

[18] Sberbank mathematically calculated the peak of the epidemic of coronavirus in Russia https://www.kxan36news.com/sberbank-mathematically-calculated-the-peak-of-the-epidemic-of-coronavirus-in-Russia

